# Lesion-Network Mapping of Post-Stroke Depressive Symptoms: Evidence from Two Prospective Ischemic Stroke Cohorts

**DOI:** 10.1101/2024.12.31.24319837

**Authors:** Ana Sofía Ríos, Uchralt Temuulen, Ahmed Khalil, Kersten Villringer, Huma Fatima Ali, Asli Akdeniz, Ulrike Grittner, Matthias Becher, Torsten Rackoll, Alexander Heinrich Nave, Pia S. Sperber, Thomas Liman, Christian Otte, Matthias Endres, Anna Kufner

## Abstract

**Background:** Post-stroke depression (PSD) affects up to one-third of stroke survivors, significantly impacting rehabilitation success and quality of life. However, its underlying pathophysiology remains unclear.

**Methods:** We analyzed two independent, prospective ischemic stroke cohorts (PROSCIS-B NCT01363856 and BAPTISe NCT01954797; total N=377) to identify brain regions and networks associated with depressive symptoms post-stroke. Lesion-symptom mapping (LSM) assessed associations between lesion location and depressive symptoms measured via the Center for Epidemiologic Studies Depression Scale (CES-D) up to 12 months post-stroke, while lesion network mapping (LNM) evaluated lesion connectivity with brain networks. We explored correlations between spatial similarity to the LNM-identified network and CES-D scores using linear regression models.

**Results:** LSM revealed no significant associations between lesion location and depressive symptoms. In contrast, LNM showed that lesion connectivity to brain regions—including the frontal pole, middle and inferior frontal gyri, inferior temporal gyrus, supramarginal gyrus, angular gyrus, frontal orbital cortex, and thalamus—correlated with CES-D scores (r=0.12, p=0.02). These regions overlapped with canonical resting-state networks, such as the frontoparietal (Dice coefficient [DC] = 0.28), salience (DC = 0.27), and default-mode networks (DC = 0.20), as well as a previously published depression circuit (DC = 0.43).

**Conclusions:** Lesion location alone was not associated with depressive symptoms post-stroke. However, lesion connectivity analysis revealed associations with brain networks, particularly the frontoparietal, salience, and default-mode networks, suggesting that disruption to these circuits may contribute to the development of PSD up to one-year post-stroke.

## Introduction

Post-stroke depression (PSD) affects up to one-third of ischemic stroke patients and negatively influences patient quality of life, rehabilitation success, and ultimately long-term functional outcome and patient mortality^1–3^. The underlying pathogenesis of PSD remains unclear although it is likely a combination of psychosocial factors as well as biological factors including the effects of lesion location^4^. A deepening of our understanding of the underlying disease process of PSD is crucial to not only identify patients at risk, but also to optimize current treatment strategies^5,6^.

The severity of post-stroke functional impairment, as well as a history of depression and female sex, are each associated with an increased risk of PSD, however, these factors do only partly explain the occurrence of PSD.^7–9^. Lesion-symptom mapping (LSM) and lesion network mapping (LNM) are neuroimaging techniques that identify brain regions associated with clinical deficits; LSM links specific lesion locations to observed behaviors, while LNM examines the functional or structural connectivity of a lesion to identify networks related to specific deficits^10,11^. Recent LSM studies have identified critical regions in the (right-hemispheric) basal ganglia^12^, insula^13,14^, and amygdala^12^ associated with depression following stroke. Moreover, symptoms associated with a focal brain lesion can also be considered in terms of how the lesion disrupts anatomically distributed – yet functionally connected – brain regions, a phenomenon referred to as diaschisis^15,16^.

On the other hand, LNM using normative resting state-functional MRI (rs-fMRI) data to investigate the connectivity of focal lesions that lead to common symptoms^11^, has been applied to successfully map the symptom circuits of several neuropsychiatric conditions from addiction to emotional regulation, mania, and depression^17–19^. Recent LNM studies investigating network correlates of depression following focal brain injury suggest the existence of a brain-wide circuit associated with the development of depression, which appears to be consistent across study populations of differing lesion aetiologies^14,20,21^. These findings indicate that brain functional connectivity is critical for understanding the mechanisms underlying the development of depressive symptoms and could aid in refining neuromodulation therapies^19^.

Still, in the context of depressive symptoms after stroke, our current understanding of the underlying pathophysiology remains incomplete. Studies with neuroimaging have reported evidence for different relevant regions and circuits in distinct populations, ranging from the basal ganglia^12,13^ to cortical regions^22^ at the lesion level. At the functional connectivity level, reports also show diverse involvement of distinct canonical networks, including the default mode-, salience-, limbic- and attention networks^23,24^ or no involvement at all^13^. The apparent discrepancy of these studies may be due to heterogeneous patient populations studied and differing methodologies applied in terms of depression assessment and LSM/LNM techniques.

Therefore, in this study we examined the neuroanatomical underpinnings of post-stroke depressive symptoms using causal mapping of the condition including large, uniformly phenotyped, and prospectively collected imaging by applying LSM and LNM pooling together two large, independent study populations of ischemic stroke patients to identify i) focal damage to specific brain structures and ii) lesion connectivity to specific regions or networks associated with depression assessed six to twelve months post-stroke.

## Materials and methods

### Subjects

Patients included in the current analysis stem from two studies, namely the prospective observational cohort PROSCIS-B (*PROSpective Cohort of Incident Stroke Berlin;* NCT01363856)^25^ and the prospective observational cohort BAPTISe (‘Biomarkers and Perfusion- Training-induced changes after Stroke; clinicaltrials.gov identifier: NCT01954797).^26^

PROSCIS-B was included patients ≥ 18 years old with first-ever ischemic stroke. Exclusion criteria included history of prior stroke, patients with brain tumor or brain metastasis, and/or participation in an intervention study. A detailed description of PROSCIS-B can be found in the previously published study protocol^25^. An MRI was not part of the primary study protocol and images were obtained retrospectively from clinical routine, wherever available. Patients from the PROSCIS-B population were selected based on the following inclusion criteria for the current analysis: 1) patients who received at least one MRI during standard clinical care following the index event 2) visible acute ischemic lesion on initial MRI and 3) available data on depression reported at one-year follow-up.

Patients were enrolled in the BAPTISe study if they had a sub-acute ischemic stroke (5–45 days following symptom onset) and a Barthel Index score of <65 at the time of screening. For the current analysis we selected patients who had an MRI with a visible ischemic lesion on the pre-intervention MRI and available data on depression reported at the 6-month follow-up appointment following enrollment. Of note, BAPTISe in contrast to PROSCIS-B, did not include first-ever stroke only. All participants provided informed consent. The studies were conducted in accordance to the Declaration of Helsinki and were approved by the local ethics committee in Berlin.

### Outcome assessment

In both patient populations, depression was assessed using a widely used screening tool called the Center for Epidemiologic Studies Depression Scale (CES-D)^27^. CES-D is a self-reported 20-item questionnaire with a maximum score of 60 points with questions involving the presence and frequency of depressive mood, feelings of guilt and worthlessness, helplessness and hopelessness, psychomotor retardation, loss of appetite, and sleep disturbance^28^. The higher the score, the more severe the symptoms of depression are considered. Based on previous literature, clinical depression was defined by a cut-off value of ≥ 16, also in patients with stroke^29,30^. Patients in the PROSCIS-B cohort were assessed for depressive symptoms at 1-year follow-up, while those in the BAPTISe cohort were assessed 6 months after study enrollment.

### Imaging

All patients included underwent a standard stroke imaging protocol on a 3-Tesla scanner which included a susceptibility-weighted or T2* sequence, diffusion-weighted imaging (DWI), “time-of-flight" MR angiography, and a fluid-attenuated inversion recovery sequence (FLAIR). Lesions were manually delineated on all slices on which the lesion was visible on DWI and FLAIR sequences using MRIcron (https://www.nitrc.org/projects/mricron) by an experienced physician (H.F.A) and supervised by at least two independent stroke imaging experts including one board-certified radiologist (A.Ku., K.V.) in PROSCIS-B. In BAPTISe, lesions were delineated by a neurologist using the semi-automated lesion masking toolbox clusterize^31^ on FLAIR sequences and supervised by an expert radiologist (K.V.). For both datasets, all lesions were delineated blinded to clinical data. Lesion masks were then co-registered to brain-extracted and bias corrected b0 images (Brain Extraction Tool by FMRIB Software Library FSL^32^) and normalised to the MNI152 space, 1 × 1 × 1 mm^33^. Normalisation was performed using the SyN algorithm in the Advanced Normalization Tools in Python (ANTsPy^34^). Lesion brain coverage was calculated by adding the lesion files of each dataset to create a single file (heat-map), this file was further binarized and multiplied by a brain mask of the brain in MNI152 space to obtain the overlapping volume (percentual coverage of the brain by the heat-map).

For each lesion, a lesion connectivity map was derived. In short, each patient lesion was used as a binary mask to calculate functional connectivity based on a normative functional connectome^35^ obtained from 1000 healthy subjects rs-fMRI scans as described in detail previously^11,36^ using the lead-mapper function from the lead-dbs software37(lead-dbs.org). In this approach, average blood oxygenation level-dependent (BOLD) signal of voxels within the lesion mask is correlated to every other voxel in the brain of the 1000 subjects in the connectome and averaged to obtain a connectivity map for each lesion mask comprising Pearson correlation coefficients, which were then Fisher-*z*-transformed.

### Statistical analysis

Statistical analyses were performed using STATA IC version 17 (StataCorp, College Station, Texas, USA). A linear regression analysis was conducted with depressive symptom severity (measured by CES-D scores, treated as a continuous variable) as the dependent variable. To identify clinical parameters independently associated with depressive symptom severity, both univariate and multivariate regression analyses were performed for CES-D scores up to 1 year post-stroke. Independent variables included age, sex (reference: female), log transformed baseline lesion volume (mL), baseline NIHSS score, and baseline cognitive impairment.

To determine which parameters should be included as covariates in the imaging analyses, we tested multiple regression models with varying combinations of covariates: baseline NIHSS score, log-transformed lesion volume, age, and sex. Model selection was based on the Akaike Information Criterion (AIC)^38^, which was calculated separately for the PROSCIS-B dataset (n=307), the BAPTISe dataset (n=70), and the pooled sample (n=377). Variables included in the best-fitting model were subsequently adjusted for in the imaging analyses as part of a sensitivity analysis. For increased sensitivity and robustness, the datasets were pooled together for further analyses.

### Lesion Symptom Mapping (LSM) of post-stroke depressive symptoms

We performed a voxel-wise and an atlas-based LSM analysis using the NiiStat software (https://www.nitrc.org/projects/niistat/) to explore the relationship between lesion location and severity of depression symptoms after stroke (continuous CES-D scores) in the pooled dataset (n=377). Minimum lesion overlap was set to 5% of the sample^10^ (damage of voxel/region in at least 19 patients). The analysis was adjusted for lesion volume to reduce the confounding effects of larger lesions^39^. The analysis was set to run for 5,000 permutations in a voxel-wise manner for robust statistical testing, controlling for family-wise error (FWE). A significance threshold was set at α=0.05 (two-sided). The same analysis was run in an atlas-based manner, using the Harvard Oxford Cortical and Subcortical Structural atlases^40^ with two aims: i) to make results more interpretable in terms of neuroanatomical regions and ii) to allow the detection of signals that might be too weak to survive a voxel-wise analysis.

### Lesion Network Mapping (LNM) of post-stroke depressive symptoms

We used a data-driven approach to identify brain regions connected to stroke lesions in patients with depressive symptoms. A permutation analysis was performed using individual lesion connectivity maps as the dependent variable and CES-D scores (centred) as the independent variable in the pooled dataset of 377 patients. The data was put into a General Linear model (GLM) matrix in FSL randomise^41^ and the analysis was run for 5000 permutations using [1] and [-1] contrasts to extract regions most associated to CES-D scores above the mean. Results were corrected for multiple comparisons using FWE, and a threshold-free cluster enhancement (TFCE) was applied to identify cluster-forming voxels within the connected regions. Note: CES-D scores were centred by subtracting the group mean from the score of each subject. Next, the FWE-corrected map of regions associated with depressive symptoms was used as a mask and input into the atlas query tool from FSL^32^ using the Harvard-Oxford Cortical and Subcortical Structural Atlases^40^, and the Diedrichsen (cerebellar) Atlas^42^ to label the regions with the strongest representation within the results- mask by obtaining a probability value for each region in the selected atlases.

To evaluate the relationship between lesion connectivity and depressive symptoms, spatial similarity (Pearson correlation) between the FWE-corrected permutation map and individual lesion connectivity maps was calculated, and these correlation coefficients were used as independent variable in a regression model with CES-D scores as dependent variable.

To assess whether the identified network generalized across datasets, we performed cross-dataset validation by calculating the spatial similarity (Pearson correlation) between the FWE-corrected map from the permutation analysis of one dataset and individual lesion connectivity maps from an unseen dataset. These correlation coefficients were then entered into a regression model to estimate CES-D scores in the unseen dataset based on spatial similarity.

Additionally, the resulting FWE-corrected map was compared to canonical rs-fMRI networks and similarity was determined by calculating a Dice coefficient (DC) between the cortical regions of the FWE-corrected results of the LNM analysis and each of the seven main (cortical) canonical networks parcellated by Yeo et al.^43^ (https://identifiers.org/neurovault.collection:1057), with the goal of determining whether disrupted connectivity to a specific network might be driving depressive symptoms after stroke. Similarly, a DC was calculated between our resulting map and the convergent depression circuit published previously by Siddiqi et al. to test whether our ischemic lesion- connectivity results were comparable to a map evaluating depression derived from distinct lesion aetiologies and neuromodulation targets^20^

(https://identifiers.org/neurovault.collection:13075). For the last calculation, the raw version of our permutation results (T-map) was used to make it comparable to the publicly available version of the convergent depression circuit.

### Sensitivity Analysis

To ensure that the results remained stable when adjusting for potential confounding effects of covariates, the LNM pipeline was repeated for the pooled dataset (n=377) accounting for relevant covariates identified in the AIC analysis (NIHSS on admission and sex; **Supplementary Table 2).** As in the unadjusted analysis, a regression model to estimate CES-D scores based on spatial similarity was performed.

### Data availability statement

Data supporting the results of this study is available upon reasonable request from the corresponding author.

### Code availability

Open source software was used for the pre-processing and analysis of the data, including: Lead-DBS (https://github.com/netstim/leaddbs), FSL 6.0.6.4 (https://fsl.fmrib.ox.ac.uk/fsl/fslwiki/) and ANTsPy (https://github.com/ANTsX/ANTsPy). The code used to perform AIC, and cross-validations has been made publicly available at https://osf.io/pgnwh/?view_only=015d9716f84d478eb6e2c5cf7d41fff3.

## Results

### Patient Characteristics and Depression Outcomes

In the total cohort (N=377), mean age was 65.4 (SD 12.8) and 35.8% were female; patient demographics in terms of age and sex across the two datasets were similar. Stroke severity however was higher in BAPTISe (median NIHSS on admission 9 IQR 6-12 versus median of 2 IQR1-4 in PROSCIS-B) with corresponding larger ischemic lesion volumes. Patients in the BAPTISe dataset exhibited a higher cerebrovascular risk profile in terms of frequency of risk factors and white matter lesion load. Stroke etiology categorized based on TOAST criteria was similar across both populations, although small vessel occlusion strokes were more common in the BAPTISe population (**Table 1**).

**Table 1.**
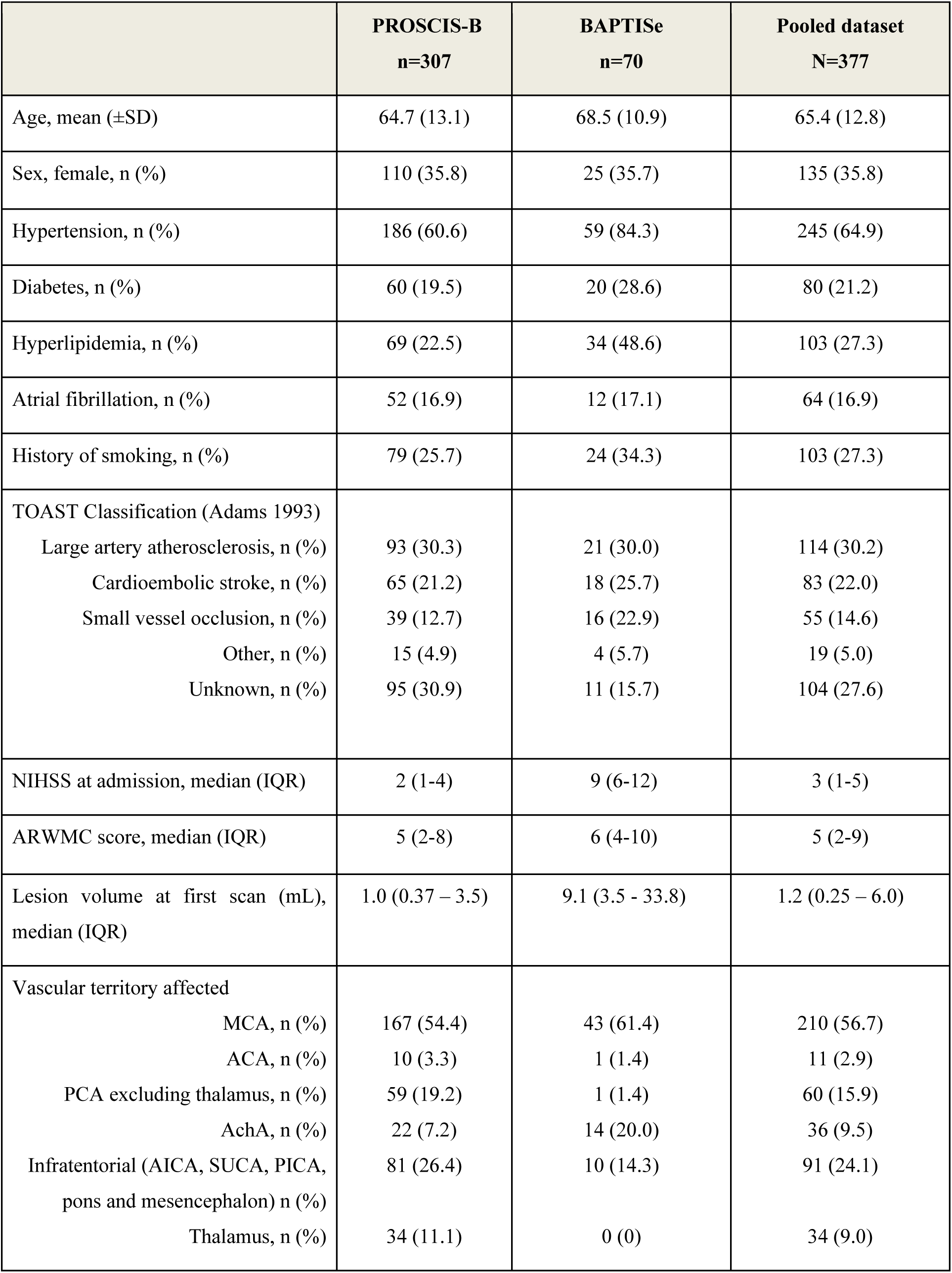
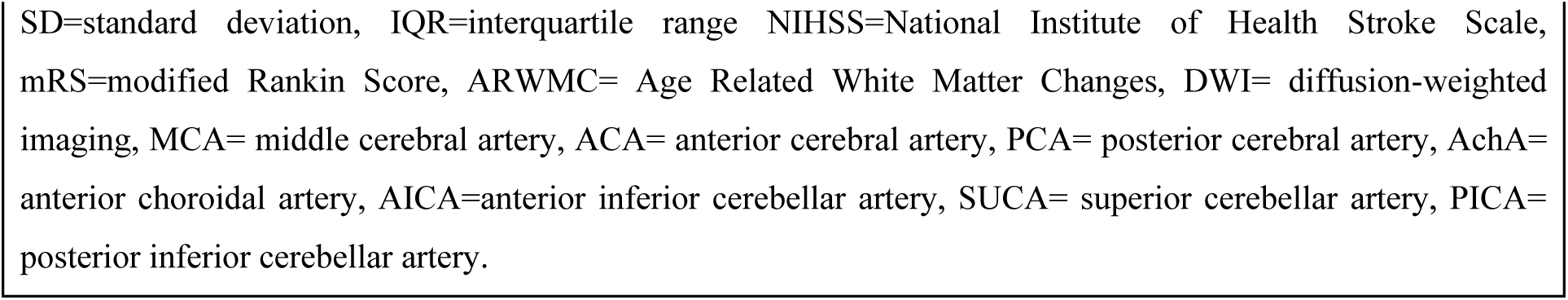
Descriptive patient demographics for each study population and pooled dataset.

|  | **PROSCIS-B**<br>**n=307** | **BAPTISe**<br>**n=70** | **Pooled dataset**<br>**N=377** |
| --- | --- | --- | --- |
| Age, mean (±SD) | 64.7 (13.1) | 68.5 (10.9) | 65.4 (12.8) |
| Sex, female, n (%) | 110 (35.8) | 25 (35.7) | 135 (35.8) |
| Hypertension, n (%) | 186 (60.6) | 59 (84.3) | 245 (64.9) |
| Diabetes, n (%) | 60 (19.5) | 20 (28.6) | 80 (21.2) |
| Hyperlipidemia, n (%) | 69 (22.5) | 34 (48.6) | 103 (27.3) |
| Atrial fibrillation, n (%) | 52 (16.9) | 12 (17.1) | 64 (16.9) |
| History of smoking, n (%) | 79 (25.7) | 24 (34.3) | 103 (27.3) |
| TOAST Classification (Adams 1993) |  |  |  |
| Large artery atherosclerosis, n (%) | 93 (30.3) | 21 (30.0) | 114 (30.2) |
| Cardioembolic stroke, n (%) | 65 (21.2) | 18 (25.7) | 83 (22.0) |
| Small vessel occlusion, n (%) | 39 (12.7) | 16 (22.9) | 55 (14.6) |
| Other, n (%) | 15 (4.9) | 4 (5.7) | 19 (5.0) |
| Unknown, n (%) | 95 (30.9) | 11 (15.7) | 104 (27.6) |
| NIHSS at admission, median (IQR) | 2 (1-4) | 9 (6-12) | 3 (1-5) |
| ARWMC score, median (IQR) | 5 (2-8) | 6 (4-10) | 5 (2-9) |
| Lesion volume at first scan (mL),<br>median (IQR) | 1.0 (0.37 – 3.5) | 9.1 (3.5 - 33.8) | 1.2 (0.25 – 6.0) |
| Vascular territory affected |  |  |  |
| MCA, n (%) | 167 (54.4) | 43 (61.4) | 210 (56.7) |
| ACA, n (%) | 10 (3.3) | 1 (1.4) | 11 (2.9) |
| PCA excluding thalamus, n (%) | 59 (19.2) | 1 (1.4) | 60 (15.9) |
| AchA, n (%) | 22 (7.2) | 14 (20.0) | 36 (9.5) |
| Infratentorial (AICA, SUCA, PICA,<br>pons and mesencephalon) n (%) | 81 (26.4) | 10 (14.3) | 91 (24.1) |
| Thalamus, n (%) | 34 (11.1) | 0 (0) | 34 (9.0) |

SD=standard deviation, IQR=interquartile range NIHSS=National Institute of Health Stroke Scale, mRS=modified Rankin Score, ARWMC= Age Related White Matter Changes, DWI= diffusion-weighted imaging, MCA= middle cerebral artery, ACA= anterior cerebral artery, PCA= posterior cerebral artery, AchA= anterior choroidal artery, AICA=anterior inferior cerebellar artery, SUCA= superior cerebellar artery, PICA= posterior inferior cerebellar artery.

In the total cohort, 97 (25.7%) reported scores above the CES-D cut-off (CES-D ≥ 16) 6 to 12 months post-stroke, indicating clinically relevant depressive symptoms. Mean depression score (CES-D) of the entire cohort was 11.0 (SD 9.6). Mean CES-D did not differ significantly between cohorts (PROSCIS-B 10.5 SD 9.4 versus BAPTISe 10.6 SD 7.0, p=0.33). In bivariate as well as multivariable regression analysis including age, sex, NIHSS on admission, lesion volume and cognitive impairment at baseline, only female sex was substantially associated with depression (adjusted coefficient: 3.9 95% CI 1.7 – 6.2, p=0.001; Supplementary Table 1).

### Lesion Location and Association with Depressive Symptoms

A heatmap of the included lesions is depicted in **Figure 1** based on CES-D scores assessed at 1 year (PROSCIS-B), and 6 months (BAPTISe). Although patients who developed clinically significant depressive symptoms (CES-D ≥ 16) had visibly higher right-hemispheric lesion coverage compared to non-depressed patients, there was no substantial difference in terms of frequency of right-hemispheric (42.5% vs. 40%) or bi-hemispheric strokes (16.4% vs. 8.7%) across groups. Lesion coverage in total was high with 82% voxels of the MNI-template affected.

**Figure 1.**
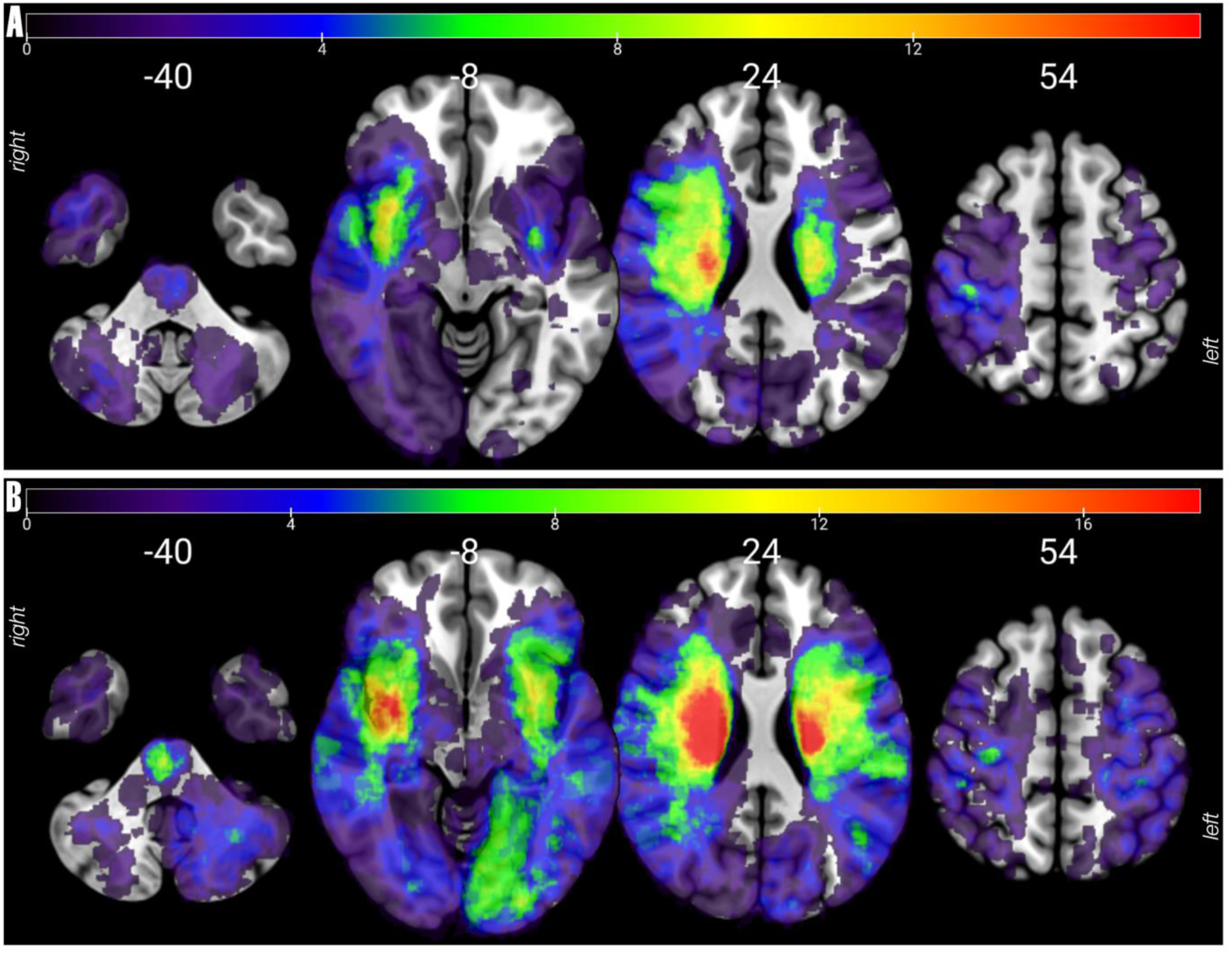
Heat-map of pooled dataset. Lesions pooling PROSCIS-B and BAPTISe (n=377) were added and overlaid on axial slices of an MNI template. A) Heat-map of lesions of patients with CES-D scores ≥ 16 (n=97), maximum overlap of 16 subjects. B) Heat-map of lesions of patients with CES-D scores < 16 (n=280), maximum overlap of 35 subjects.

Voxel-wise LSM did not yield any significant associations between lesion location and depressive symptoms evaluated by continuous CES-D scores at follow up. Atlas-based LSM (using the Harvard Oxford Cortical and Subcortical Structural atlases(Desikan et al., 2006; Makris et al., 2006) revealed no statistically significant positive associations (p<0.05, z>3.62) with depressive symptoms, however, the regions that showed highest z values (top 30%) include right-hemispheric: amygdala (z=3.2), nucleus accumbens (z=3.2), lateral occipital cortex (z=3.0), temporal pole (z=2.7), middle frontal gyrus (z=2.5), thalamus (z=2.5), inferior frontal gyrus (pars opercularis, z=2.4), angular gyrus (z=2.4) and occipital pole (z=2.4; **Supplementary** Figure 1).

### Functional Connectivity Correlates of Depressive Symptoms

The LNM analysis of depressive symptoms in the pooled dataset revealed connectivity to the following regions: frontal pole, insular cortex, middle frontal gyrus, inferior frontal gyrus (pars opercularis and triangularis), temporal pole, superior temporal gyrus (posterior division) middle temporal gyrus (temporooccipital part and posterior division), inferior temporal gyrus (posterior division), supramarginal gyrus (posterior division), angular gyrus, cingulate gyrus (posterior division), precuneus cortex and frontal orbital cortex and right thalamus, 1-pFWE values ranged from 0 to 0.61. Although connectivity to these regions is observed in both hemispheres, the spread of the map is higher on the right hemisphere (**Figure 2**, **Figure 3**). For a complete list of connected regions, refer to **Supplementary Table 3**. The regression model showed a modest correlation of r = 0.12 (p = 0.02, **Figure 2**), suggesting a subtle association between the regions identified in the permutation analysis and depressive symptoms measured by CES-D scores. Similarly, sensitivity analysis (adjusted for NIHSS on admission and sex) showed a positive association between lesion connectivity to the frontal pole, middle frontal gyrus, inferior frontal gyrus (pars opercularis), middle temporal gyrus (temporooccipital part), supramarginal gyrus, angular gyrus and precuneus cortex to depressive symptoms after stroke with a Pearsons correlation coefficient of r=0.11 (p=0.04; **Supplementary** Figure 2).

**Figure 2.**
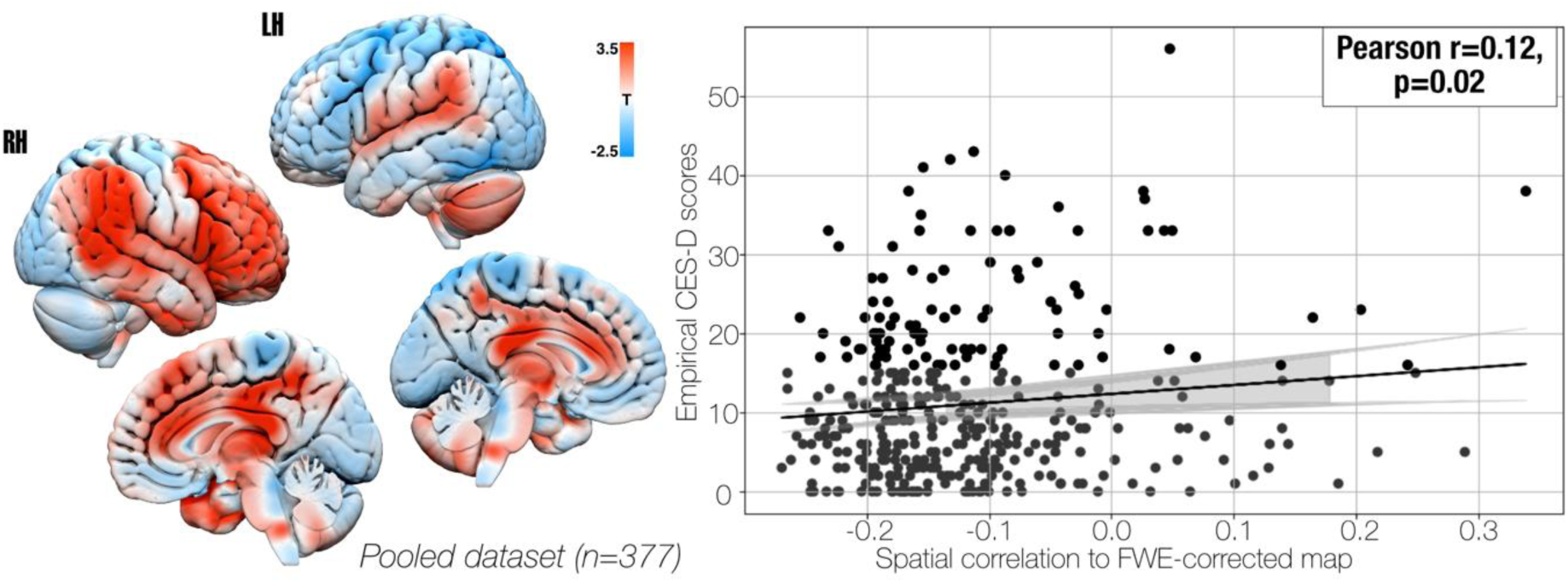
LNM of post-stroke depression. Left: statistical maps representing positive (white-red) and negative (white-blue) associations of lesion connectivity and depressive symptoms after stroke based on demeaned CES-D scores overlaid on a 3D brain surface in MNI space, output from randomise was used in form of T-maps to facilitate visualization. Right: the scatter plot shows the relationship between spatial correlations (x-axis) and empirical CES-D scores (y-axis) in the pooled dataset. Each data point represents one patient. The fitted regression line (shown in black) estimating CES-D scores (r=0.12, p=0.02) from spatial correlations of the LNM resulting map to each lesion connectivity map within the sample, shaded grey area indicating the 95% confidence interval of the fitted model. RH: Right hemisphere, LH: Left hemisphere.

**Figure 3.**
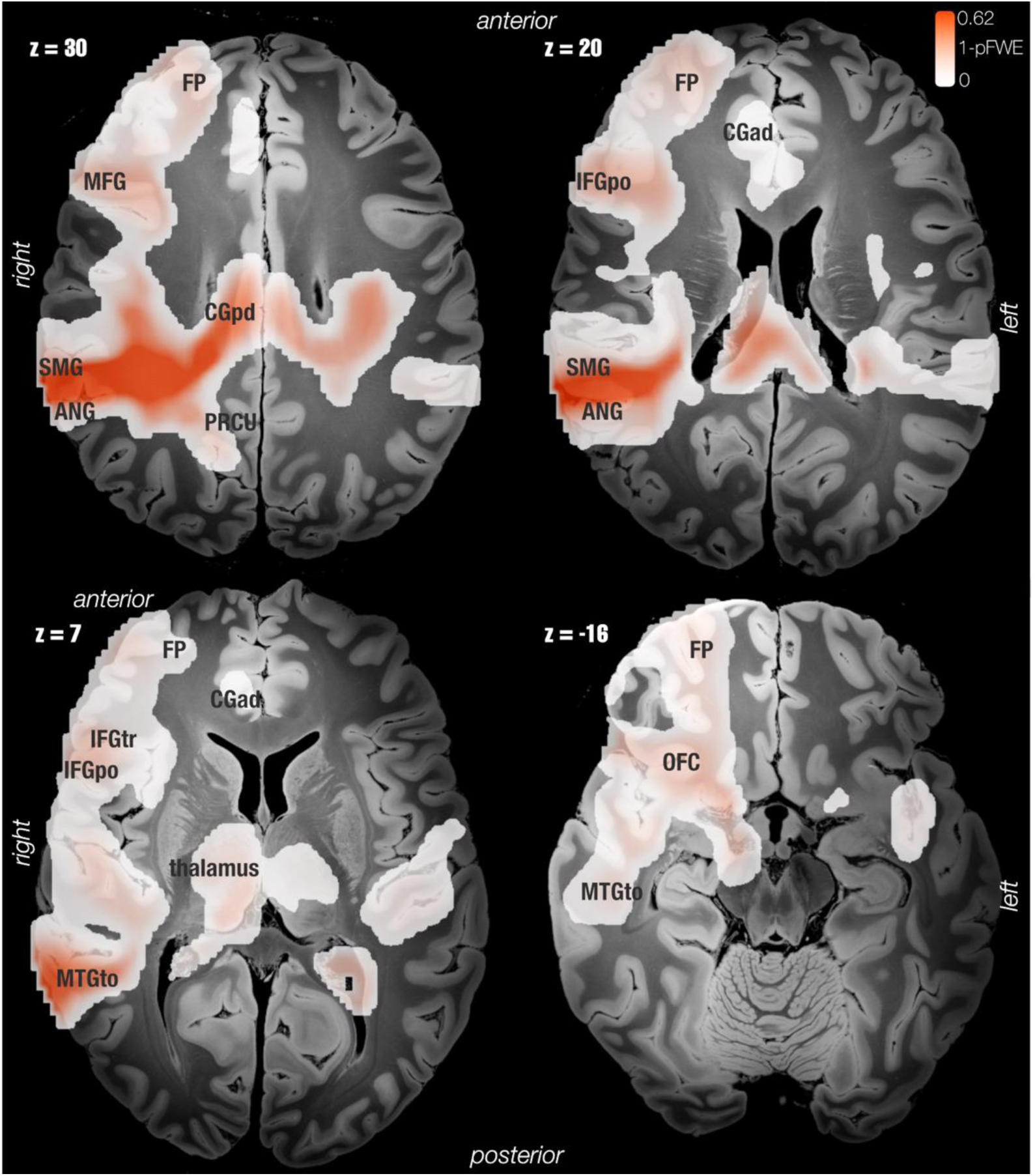
**Regions associated to depressive symptoms in pooled dataset**. FWE-corrected map of positive associations to depressive symptoms based on demeaned CES-D scores in the pooled dataset (n=377) overlaid on axial slices of an ex-vivo MNI space template acquired at 7T with 100µm resolution. The color bar indicates corrected 1-pFWE values, ranging from 0 (white) to 0.62 (red), lesion connectivity to the following regions: frontal pole (FP), insular cortex, middle frontal gyrus (MFG), inferior frontal gyrus pars opercularis (IFGpo) and pars triangularis (IFGtr), temporal pole, superior temporal gyrus (posterior division), middle temporal gyrus (temporooccipital part, MTGto) and posterior division, inferior temporal gyrus (posterior division), supramarginal gyrus (posterior division, SMG), angular gyrus (ANG), cingulate gyrus (posterior division, CGpd), precuneus cortex (PRCU), frontal orbital cortex (OFC), and right thalamus, was associated to depression symptoms (CES-D scores) based on LNM permutation analysis.

The cross-dataset validation using linear regression analysis revealed no significant correlations between lesion connectivity maps and CES-D scores, with Pearson correlation coefficients of r=0.13 (p=0.27) for BAPTISe using the PROSCIS-B map, and r=-0.01 (p=0.81) for PROSCIS-B using the BAPTISe map (**Supplementary** Figure 3). For details on the regions identified in the LNM analysis for each dataset separately, refer to **Supplementary Tables 4 and 5**.

Dice coefficient (DC) calculations assessing the spatial similarity between LNM results of the pooled dataset and rs-fMRI networks^43^ (https://identifiers.org/neurovault.collection:1057) revealed low-to-moderate overlap with the frontoparietal network (DC = 0.38), salience network (DC = 0.27) and default mode network (DC = 0.20), as for the limbic network (DC = 0.18), and somatomotor network (DC = 0.17) low overlap was observed (**Figure 4**). No overlap was observed for the visual network. LNM results of the pooled dataset showed a moderate overlap (DC = 0.43) with the convergent depression circuit^20^, specifically in regions such as the frontal pole, insular cortex, middle and inferior frontal gyri, precentral gyrus, supramarginal and angular gyri, precuneus cortex, frontal orbital cortex, central opercular

**Figure 4.**
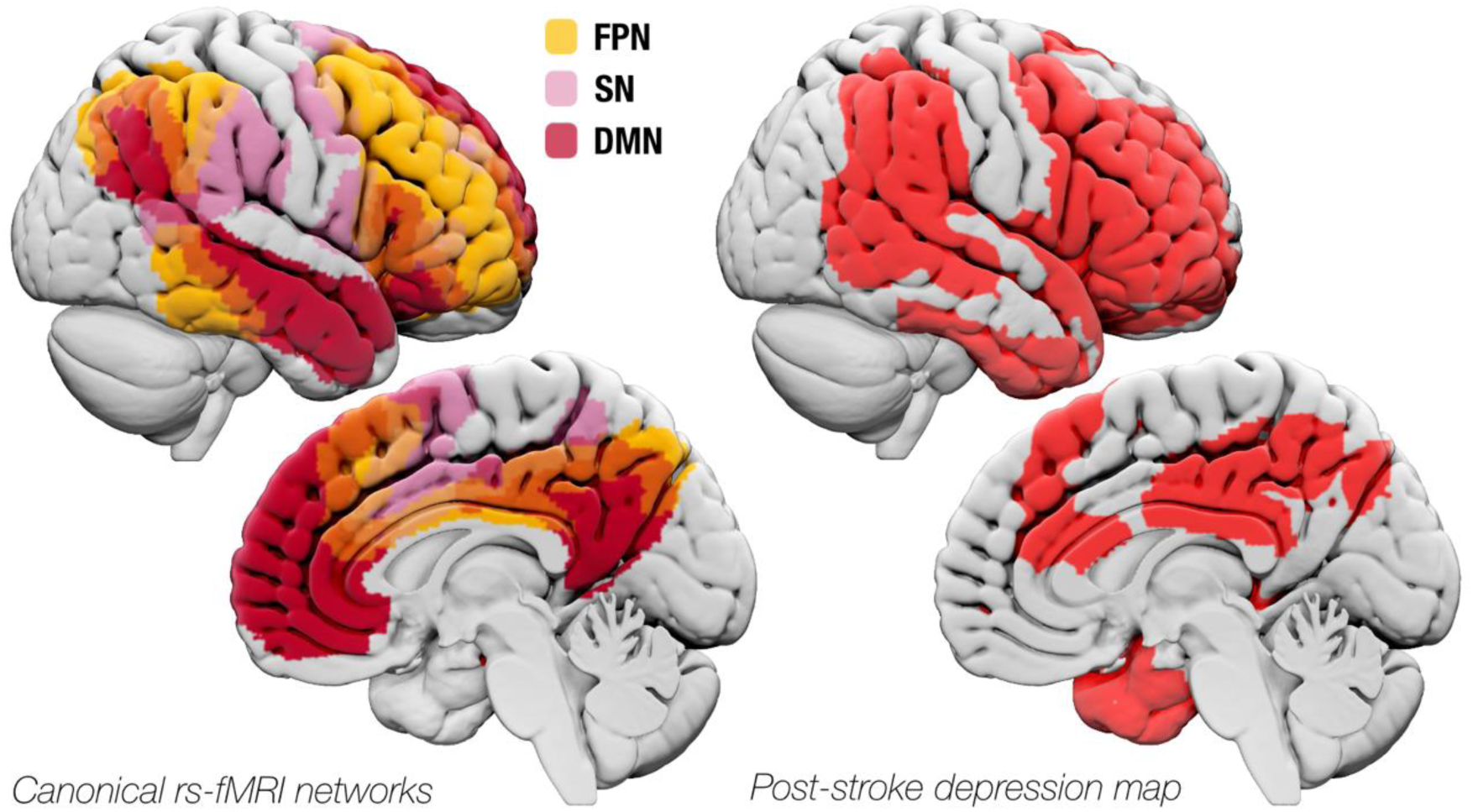
**Comparison of LNM results to canonical rs-fMRI networks**. The left panel shows canonical resting-state fMRI (rs-fMRI) networks(Yeo et al., 2011), including the frontoparietal network (FPN, yellow), salience network (SN, pink) and default mode network (DMN, red). The right panel illustrates the LNM results from the pooled dataset (n=377), highlighting cortical regions associated with depression following stroke (in red).

cortex, and parietal operculum cortex (**Figure 5**).

**Figure 5.**
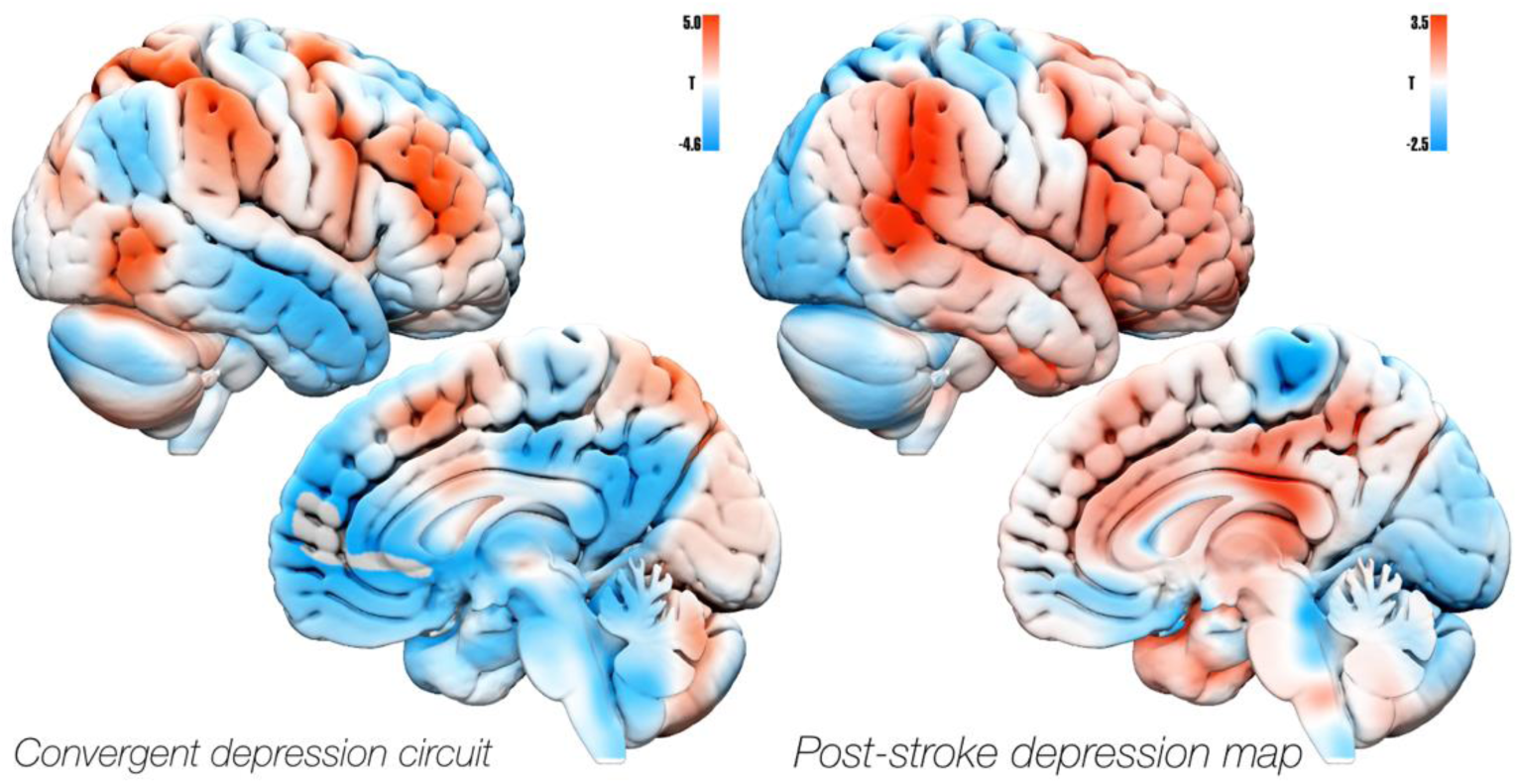
**Comparison of PSD map to published depression circuit**. The post-stroke depression map (right) was compared with the convergent depression circuit from Siddiqi et al. (left)^20^. The maps showed moderate overlap based on Dice coefficient (DC=0.43). The overlapping regions include: the frontal pole, insular cortex, middle and inferior frontal gyri, precentral gyrus, supramarginal gyrus, angular gyrus, precuneus cortex, frontal orbital cortex,central opercular cortex, and parietal operculum cortex. White-red color indicate positive-, while white-blue colors indicate negative associations, with the respective T-values indicated by the color bars.

## Discussion

In this study, we investigated the neuroanatomical substrates of post-stroke depressive symptoms in 377 subjects from two independent stroke cohorts, with depressive symptoms assessed using the CES-D at 6- and 12-months post-stroke. Focal damage to specific brain regions was not significantly associated with depressive symptoms. However, lesion network mapping revealed that lesion connectivity to a brain circuit, including several cortical regions and the right thalamus, was associated with severity of depressive symptoms. These findings provide further evidence that network dysfunction, rather than isolated focal damage, may contribute to the development of depressive symptoms after stroke Patients from the PROSCIS-B dataset are representative of a mild-to-moderate stroke population in terms of stroke aetiology, demographics, and risk factors^44,45^ (**Table 1),** whereas patients included in BAPTISe represent a more severely affected stroke population with a higher cerebrovascular risk profile and larger lesions. In multivariate regression analysis within the pooled dataset, only female sex was identified as independent predictor of higher depression scores (**Supplementary Table 1**), which stands in line with the literature^7–9^. Overall, lesion coverage in the current study was high (82%) compared to previous PSD-LSM and LNM studies^12–14,22^, and infratentorial regions and the thalamus were well represented, with 26% and 11%, respectively (**Figure 1)**.

In terms of lesion location, we did not find any statistically significant positive associations in our dataset in a voxel-wise nor atlas-based LSM analysis. Previous studies have reported positive LSM results for PSD and found that focal damage to specific brain structures such as right dlPFC and inferior frontal gyrus^22^, as well as basal ganglia^12,13^, and amygdala^12^ was associated with depression. The absence of significant associations in our study may reflect the heterogeneity of post-stroke depressive symptoms, which may not be fully captured by the CES-D score as it primarily assesses general depressive symptomatology and does not discriminate modalities of negative affect. This lack of specificity may limit its ability to identify a single lesion or brain region uniquely associated with this symptomatology. Nevertheless, we observed numerically positive associations in certain previously reported structures, including the amygdala, middle, and inferior frontal gyri, although these did not reach statistical significance (**Supplementary** Figure 1).

When exploring lesion connectivity contributions to depressive symptoms post-stroke, the LNM analysis revealed positive associations between CES-D scores and lesion connectivity to the frontal pole, middle frontal gyrus, inferior frontal gyrus (pars opercularis), temporal pole, middle temporal gyrus (temporooccipital part), inferior temporal gyrus (posterior division), supramarginal gyrus (posterior division), angular gyrus, cingulate gyrus (posterior division), precuneus cortex, and frontal orbital cortex, as well as the right thalamus (**Figures 2 and 3, Supplementary Table 3**). The resulting map was used to estimate CES-D scores based on spatial correlation to the lesion connectivity map of each subject within the dataset (r=0.12, p=0.02, **Figure 2**), showing a small, but statistically significant effect. However, cross-dataset validation revealed no significant correlations between lesion connectivity maps and CES-D scores, indicating that the observed associations may not generalize across independent datasets (**Supplementary** Figure 3). The LNM findings in the pooled dataset remained consistent when adjusting for NIHSS and lesion volume (**Supplementary** Figure 2).

The functionally connected structures identified through LNM, showed relevant overlap with three described canonical resting-state fMRI (rs-fMRI) networks (Yeo^43^): the *frontoparietal network* (FPN, DC=0.38), *salience network* (SN, DC=0.27), and *default mode network* (DMN, DC=0.20) compared to other canonical networks (**Figure 4**). Interestingly, the involvement of these networks in both major depressive disorder (MDD) and PSD has been previously described^14,19,20^. Overall, dysfunction in these networks, specifically as an imbalance in connectivity between them, was described as a potential causal mechanism underlying impairments in flexible cognition and executive function, along with difficulty transitioning from introspective to externally-focused tasks^46^.

In short, the FPN is involved in cognitive control, decision-making, and working memory^47^. In the context of depression, a hypoactive FPN could explain deficits in cognitive function^46^. The SN, on the other hand, plays a key role in attentional switching between cognitive states, such as self-monitoring processes, and high task involvement -- in other words, alternating between internally focused networks like the DMN, and task-associated networks like the FPN and the central executive network^48,49^. Furthermore, DMN activity is engaged in self-referential thinking, mind-wandering, episodic memory retrieval, and social cognition^50^. Increased DMN activation was previously described in depressed patients, specifically linked to symptoms such as rumination or an overrepresentation of negative, self-referential thoughts^51^.

Following the concept of network dysfunction, Siddiqi et al. advanced the discussion by describing a common depression circuit derived from focal lesions, non-invasive and invasive stimulation targets^20^. This convergent depression circuit was most like the dorsal attention network and FPN and highlighted the advantage of network mapping methodologies to inform neuromodulation therapies. The regions associated to depressive symptoms identified in our dataset show moderate overlap (DC=0.43, **Figure 5**) to that described by Siddiqi et al. at the level of frontoparietal cortices, including the middle and inferior frontal-, supramarginal and angular gyrus and frontal orbital cortex. Our findings suggest that network disruption might contribute to the pathological changes that lead to the development of depressive symptoms after stroke, however with the data presented in this work, we were not able to validate this association across datasets.

In summary, LSM and LNM studies, including the present one, have identified various regions and networks associated to the development of depressive symptoms across diverse patient populations. Notably, we see that lesions effecting integral regions of selected canonical networks (FPN, SN, DMN) are consistently associated with the presence of depressive symptoms^12–14,20,22^. In other words, lesions may lead to a dysfunction or disbalance across these crucial networks and increase patient risk for developing depressive symptoms. In this context, it is important to highlight, that depression is a complex syndrome that involves a wide range of symptomatology spanning from affective to somatic symptoms and cognitive deficits, and that a more detailed assessment of symptom-subdomains may ultimately lead to the identification of more specific regions and networks^22^. Independent, larger, cohort studies with uniform evaluation of depression severity and specific depressive symptoms are warranted to validate our findings.

### Limitations

This study has some important limitations that warrant discussion. From a clinical point of view, we have no data available on patient history of depression or other psychiatric illnesses, therefore the presence of these pre-existing conditions could not be accounted for. Moreover, self-reported depressive symptoms were used in this study, which, although convenient for large-scale data collection, may not completely capture the multidimensional nature of depression or account for comorbid conditions that influence symptom reporting, hence, CES-D is only suitable as a screening tool for depression and cannot be used to formally diagnose PSD. It is also noteworthy that CES-D does not evaluate cognitive deficits, which are known symptoms of depression^27^. On the other hand, such screening tools are commonly used in large, prospective, stroke cohorts which increases the potential comparability of our results. Also noteworthy, is that clinical depression is often episodic in nature and is known to fluctuate over time, and assessment at one time point only may miss cases; however, the literature shows that PSD frequency is highest in the first year, remaining constant during this period^6^. In addition, we could not adjust for the potential influence of new medications taken i.e. whether patients received new antidepressant medication thus influencing depressive symptoms at 1-year follow-up. This is a notable limitation of the current study that should be accounted for in further validation studies. Furthermore, a more detailed assessment of depressive symptoms i.e. subdomains of depression that are not captured in the CES-D is warranted.

Another limitation is the potential for selection bias and attrition bias; since only patients with available MRI, lesion data, and CES-D follow-up data were considered, this subset of participants may deviate from the initial study protocols and may not fully represent the broader study population. Moreover, in the neuroimaging context, the analyses in this study primarily focused on linear relationships, which may not adequately capture the complexity of depressive symptoms. Furthermore, LNM using a normative connectome only provides an indirect measure of functional diaschisis and therefore can only explain symptoms with reduced variance compared to direct measures of functional connectivity. By using normative data from young healthy subjects - age range from 18 to 35 years - we cannot account for age-related changes in functional connectivity or individual compensation mechanisms that might occur following ischemic stroke. Nonetheless, the advantage of using this type of methodology is that it allows us to analyze large amounts of lesion data from patients coming directly from a clinical setting in which structural MRI is more commonly acquired than functional MRI. An important validation step would be to perform a similar analysis in a sub- group of patients that also undergo rs-fMRI for comparability of results.

## Conclusion

One of the main findings of this study is that lesion location alone was not associated with CES-D scores meassured up to 1-year post-stroke in our pooled analysis of 377 ischemic stroke patients. This study contributes to the current knowledge on the underlying functional “disconnections” that might lead to depressive symptoms following ischemic stroke. Although PSD is a complex, multifactorial disorder, patients with ischemic lesions functionally connected to crucial anatomical regions, part of the *frontoparietal, salience and default mode network* had a higher risk of developing depressive symptoms up to one-year after stroke. However, the inability to validate these results in independent datasets shows that larger samples and more sophisticated diagnostic tools as well as neuroimaging analytical approaches for the study of depressive symptoms after stroke are required to further investigate functional and neuroanatomical associations of PSD.

## Data Availability

Data supporting the results of this study is available upon reasonable request from the corresponding author. Code availability: Open source software was used for the pre-processing and analysis of the data, including: Lead-DBS (https://github.com/netstim/leaddbs), FSL 6.0.6.4 (https://fsl.fmrib.ox.ac.uk/fsl/fslwiki/) and ANTsPy (https://github.com/ANTsX/ANTsPy). The code used to perform AIC, and cross-validations has been made publicly available at https://osf.io/pgnwh/?view_only=015d9716f84d478eb6e2c5cf7d41fff3.

## Acknowledgements

We thank the patients who generously agreed to participate in both studies, and the Netstim Lab for kindly offering methodological support. Especially we would like to thank Dr. Bassam Al-Fatly and Prof. Dr. Andreas Horn for their methodological support and analysis advice. We acknowledge the use of OpenAI’s ChatGPT-4 for assisting in writing and correcting the code used in this study. The tool was used to enhance code efficiency and troubleshoot errors during the analysis. All scripts and generated outputs were critically reviewed and validated by the authors to ensure their accuracy and suitability for the research objectives.

## Author contribution statement

ASR and AKu designed the study and prepared the manuscript. HFA delineated lesions from PROSCIS-B. KV delineated lesions from BAPTISe. Aku and KV supervised lesion delineation of PROSCIS-B. TU performed the pre-processing of the imaging data. AKu performed regression analysis on demographic and clinical data. ASR ran LSM and LNM analyses. AK, KV, UG, AA, MB, CO, ME participated in study design and interpretation of results. TR, AHN, PS, TL collected the data. AKu and ME jointly supervised this work.

